# Electrophysiology in the time of coronavirus: coping with the great wave

**DOI:** 10.1101/2020.03.30.20044776

**Authors:** Jia Li, Patrizio Mazzone, Lisa WM Leung, Weiqian Lin, Giuseppe D’Angelo, Jun Ma, Jin Li, Zaki Akhtar, Yuechun Li, Paolo E. Della Bella, Jiafeng Lin, Mark M Gallagher

## Abstract

**Aims:** To chart the effect of the COVID-19 pandemic on the activity of interventional electrophysiology services in affected regions.

**Methods:** We reviewed the electrophysiology laboratory records in 3 affected cities: Wenzhou in China, Milan in Italy and London, United Kingdom. We interviewed electrophysiologists in each centre to gather information on the impact of the pandemic on working patterns and on the health of staff members.

**Results:** There was a striking decline in interventional electrophysiology activity in each of the centres. The decline occurred within a week of the recognition of widespread community transmission of the virus in each region and shows a striking correlation with the national figures for new diagnoses of COVID-19 in each case. During the period of restriction, work-flow dropped to <5% of normal, consisting of emergency cases only. In 2 of 3 centres, electrophysiologists were redeployed to perform emergency work outside electrophysiology. Among the centres studied, only Wenzhou has seen a recovery from the restrictions in activity. Following an intense nationwide program of public health interventions, local transmission of COVID-19 ceased to be detectable after February 18^th^ allowing the electrophysiology service to resume with a strict testing regime for all patients.

**Conclusion:** Interventional electrophysiology is vulnerable to closure in times of great social difficulty including the COVID-19 pandemic. Intense public health intervention can permit suppression of local disease transmission allowing resumption of some normal activity.

**CONDENSED ABSTRACT:** COVID-19 has affected every aspect of life worldwide. In the electrophysiology labs of Wenzhou, Milan and London, activity was suspended as the disease took hold. Only Wenzhou has resumed normal services, facilitated by a monumental nationwide program of public health interventions and supported by stringent testing protocols.

**WHAT’S NEW:**

- We describe the impact of the COVID-19 pandemic on interventional electrophysiology units in 3 cities: Wenzhou, Milan and London.
- In all cases, the routine work of the electrophysiology was virtually suspended within a week of the recognition of widespread virus transmission in the area.
- During the period of restricted activity imposed by the pandemic, centres have dealt with a small number of emergency ablations only, a balanced mix of atrial, ventricular and junctional arrhythmias.
- In 2 of the 3 centres, electrophysiologists were redeployed to perform other medical duties including in COVID-19 wards.
- COVID-19 infection occurred in medical and nursaing staff in 2 of the 3 centres.
- Only in the cases of Wenzhou, China, has a resumption of normal activity been possible; this follows intensive public health intervention and is protected by stringent testing.

**FUNDING:** None

**ETHICAL APPROVAL:** None required from the Research Ethics Committee (REC) London according to the type of study. Institutional ethical approval obtained at the centres of: St. George’s Hospital NHS Foundation Trust, London, UK; Local Health Authority Ethical Approval was obtained in: The Second Affiliated Hospital and Yuying Children’s Hospital of Wenzhou Medical University in Wenzhou, PR China and San Raffaele in Milan, Italy.

**CONSENT:** Informed consent was obtained from all participants/interviewees who took part in this study.

## BACKGROUND

The COVID-19 pandemic has caused suffering and death across the world since late 2019.^1,2^ Physicians and journalists encountering the arrival of the epidemic in an area typically describe a wave, a tide or a tsunami of disease. It has had a widely publicised impact on the economy of afflicted areas. Medical services have suffered massive disruption. Even services far removed from respiratory medicine and critical care have suffered because of the diversion of resources needed to support the victims of the epidemic and through the illness of staff. As a resource-intensive speciality that deals predominantly with non-emergency cases, interventional electrophysiology is particularly vulnerable to disruption.

While the pandemic disrupts the delivery of routine electrophysiology services, COVID-19 is associated with cardiac complications which could bring an additional burden of acute problems to electrophysiology.^3-5^ The relative importance of the reduction in elective cases and any increase in emergency work is undefined.

Wenzhou, Milan and London are each cities with a metropolitan area population of more than 4 million people; each has each been struck by the pandemic. During the initial months of 2020, Wenzhou experienced the greatest concentration of COVID-19 cases of any Chinese city outside the Hubei province.^6^ Following an intense program of public health interventions^7^, new cases declined and have now vanished. As of March 27^th^ 2020, Wenzhou has had no new case of COVID-19 for the last 38 consecutive days. With this, many sectors have begun to return toward normality with stringent precautions. Milan was close to the centre of the Italian outbreak of COVA-19 and had to suspend all activity the first week in March.^8^ London, like the rest of the United Kingdom has seen an abrupt rise in the disease incidence only since the second week of March 2020.^9^ The disruption for cardiac electrophysiology labs has begun.

## METHODS

We reviewed the catheter lab records of electrophysiology laboratories in a centre that experienced a high burden of COVID-19 early and near the origin of the pandemic and one in which the onset was delayed. The impact on workflow resulting from the redirection of staff and resources was correlated with the case-load of COVID-19. We charted the burden of emergency procedures performed to look for evidence of any augmentation of these arising from COVID-19, and we interviewed the front-line cardiologists for information about arrhythmic complications encountered in the COVID-19 population. We looked for instances of COVID-19 infection acquired in hospital by electrophysiology patients and staff. We documented the protocols used to permit the resumption of activity after the first wave of the epidemic and examined the success of anti-infective precautions.

## RESULTS

The first diagnosis of COVID-19 in Wenzhou Medical University was in mid-January 2020; In Lomardy a large outbreak became evident on February 22nd, while in St George’s Hospital, London cases began to arrive in large number from the second week in March. In all cases, there was a sharp downturn in activity within days after the recognition of widespread COVA-19 transmission in the area (figure 1).

**Figure 1:**
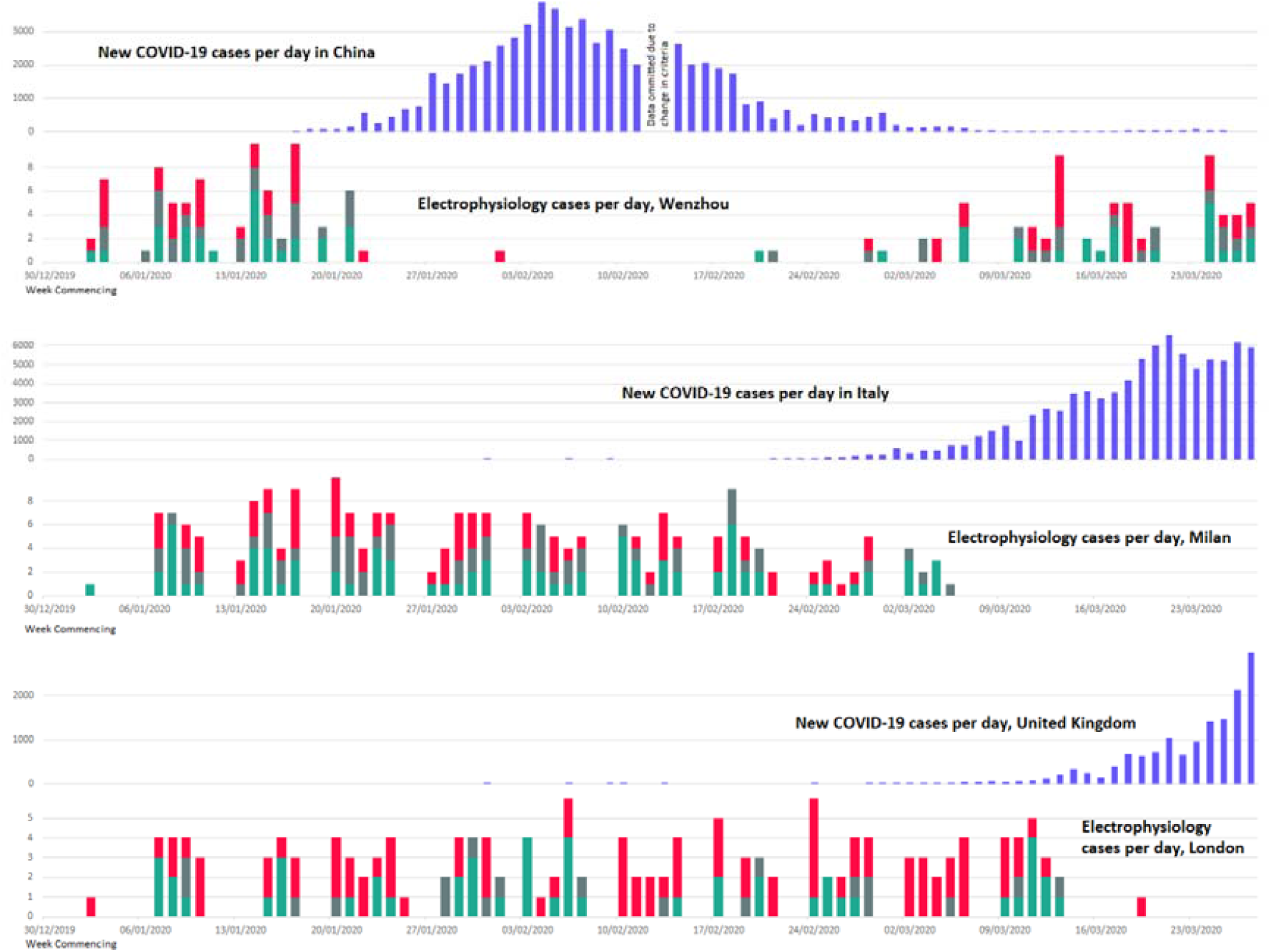
Chronology of events in the first 87 days of 2020 In each of the electrophysiology labs, routine activity was cancelled as soon as the country detected significant numbers of COVID-19 cases. During the period of suspension of routine activity, very few emergency procedures were performed. Of the 3 centres, only Wenzhou had been able to resume normal activity.

### Effect on routine work

In each of the centres, routine activity of the EP lab was suspended within a week of the first locally diagnosed COVID-19 case in the hospital (table 1). The primary reason for the prompt suspension in each case was the concern that continuing to admit patients for elective procedures would expose patients to the risk of infection from undiagnosed COVID-19 patients already in the hospital.

**Table 1:** Effects of COVID-19 on electrophysiology staff and services.

|  | Wenzhou | Milan | London |
| --- | --- | --- | --- |
| Cessation of routine ablations | yes | yes | yes |
| Redeployment of electrophysiologists |  |  |  |
| non-EP acute cardiology | yes | yes | yes |
| intensive care | no | no | yes |
| COVID-19 ward, non-intensive | yes | no | yes |
| Nosocomial COVID-19 in electrophysiology |  |  |  |
| patients infected | 0 | 2 | 1 |
| electrophysiologists infected | 0/5 | 0/10 | 2/9 |
| other department staff infected | 0 | 3 nurses | 0 |

### Electrophysiological emergencies during the COVID-19 crisis

We encountered no instance of an arrhythmia occurring as a definable consequence of COVID-19 that required ablation as an emergency procedure. A small number of patients required emergency ablation during the period of restricted activity (table 2, figure 2), but this represented less than 5% of the normal workload of the centres. The distribution of cases was similar to that encountered in normal times with supraventricular tachycardia, ventricular tachycardia and atrial fibrillation all represented.

**Table 2:** Effects of the pandemic on electrophysiology activity.

|  | Wenzhou | Milan | London |
| --- | --- | --- | --- |
| Period of restricted activity: | Jan 23<sup>rd</sup> to<br>March 2<sup>nd</sup><br>2020 | From 5<sup>th</sup><br>March 2020<br>(ongoing) | From 16<sup>th</sup><br>March 2020<br>(ongoing) |
| ablations performed during this period: |  |  |  |
| supraventricular tachycardia | 2 | 0 | 0 |
| ventricular tachycardia / ectopy | 2 | 1 | 0 |
| atrial tachycardia / fibrillation | 3 | 0 | 1 |
| Arrhythmias ablated in COVID-19 patients | 0 | 0 | 0 |

**Figure 2:**
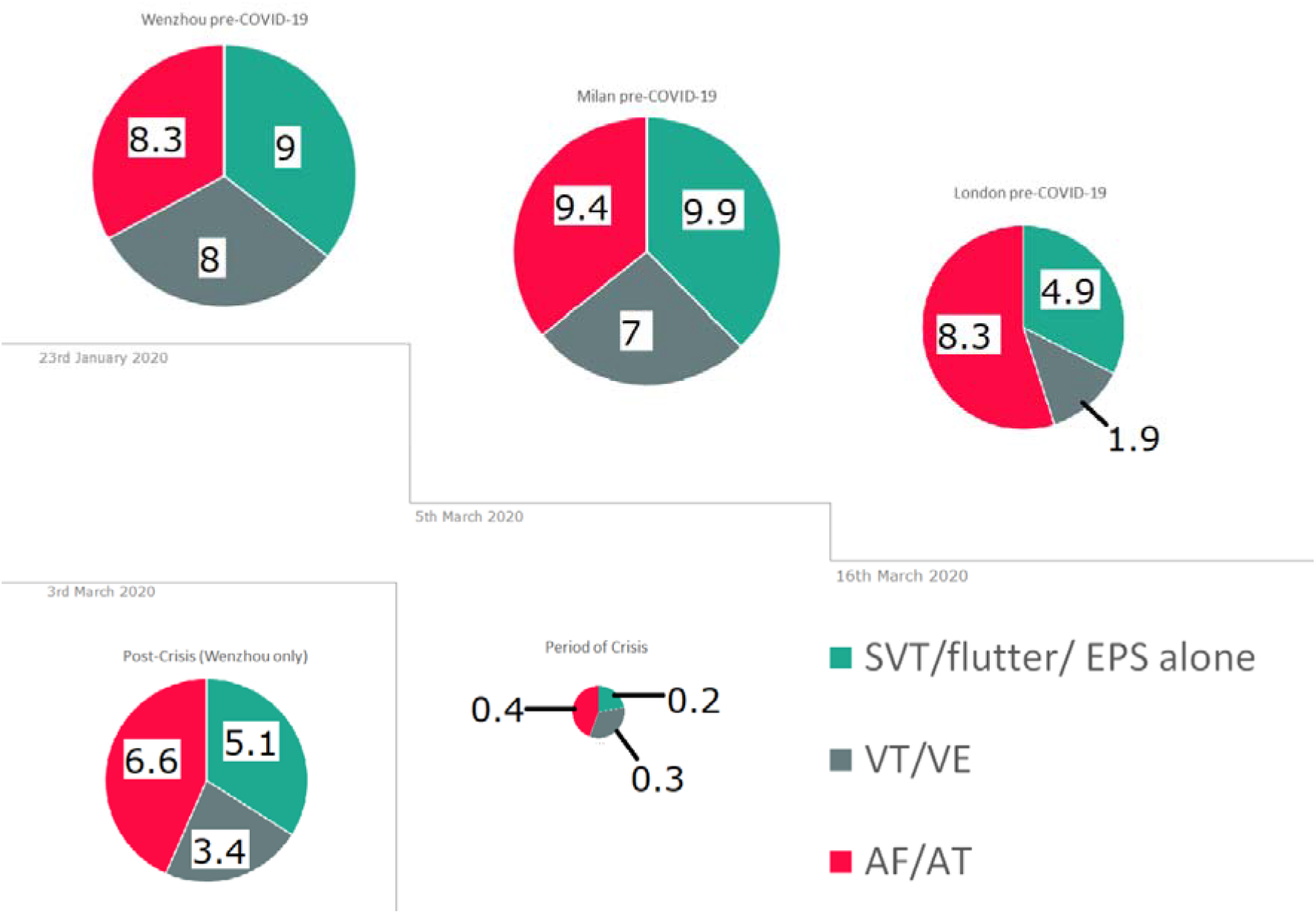
Impact of the COVID-19 epidemic on workflow in electrophysiology. The pie-charts indicate the mean number of each category of each procedure carried out per week in 2020 in each centre in the period before COVID-19 arrived, in the period of restricted activity and in the case of Wenzhou after the resumption of normal activity. The workload during the period of crisis represents the mean number of cases per week per centre across all 3 centres, and represents a fall of more than 95% compared to pre-crisis activity without a clear-cut alteration in the breakdown of procedure types.

### Nosocomial Infection

One patient in London acquired COVID-19, apparently in hospital after her ablation. She died of pneumonia associated with the condition at 18 days after the ablation.

Two electrophysiologists in London including a co-author of this paper acquired it and recovered without complication. Three nurses in a cardiology ward in Milan were infected, but none of the electrophysiologists. In Wenzhou, no staff member was infected.

### Resumption of normal activity

Of the centres involved, only Wenzhou has been able to resume routine activity. This became possible because of a country-wide suppression of the epidemic produced by an intensive program of public health interventions across the country. This included a period of lock-down accompanied by intensive contact-tracing and use of isolation. Mobile technology played an integral part. Electronic tracking of each individual was conducted with a traffic-light system to inform the person and those in close proximity of their health and travel status.

Having achieved the virtual elimination of virus transmission in most of the country by late February, routine activity resumed with stringent precautions:

1. Patients underwent a pre-assesment visit the day before hospital admission. At this visit, a nasal swab was analysed by PCR for evidence of the virus.
2. Patients with a satisfactory screening visit were admitted on the morning of the scheduled procedure and underwent a second nasal swab. If the PCR was negative for the virus, the procedure went ahead as planned.

### Outpatient activity

In all centres, outpatient activity was moved to a telephone-based or internet-based format in all possible cases from the time that widespread local transmission of the virus was recognised. Only those who were clinically urgent and were unable to to use a telephone or required an instrumental test were allowed to keep their physical appointments. Outpatient investigations were cancelled unless clinically urgent. Patients who had access to mobile-phone associated ECG recording were encouraged to use these and transmit recordings in preparation for telephone consultations.

### EP Education

As part of social distancing, all EP educational meetings were suspended in all centres, and all major international meetings have been cancelled. Electronic learning has been put into practice across several medical disciplines and improving web-based educational tools. Major medical examinations such as the Membership exam (MRCP) and university entrance exams have been delayed indefinitely.

### EP team redeployment

The EP team in Wenzhou and that in London defaulted to emergency mode at the arrival of the first COVID cases. With elective work being cancelled, the electrophysiologists concentrated on urgent inpatient intervention including pacemaker implantation to minimist the time that any patient had to spend in hospital. The centres differed in their policies on redistributing staff to other duties.

## DISCUSSION

> “*I think it better that in times like these*
>
> *A poet’s mouth be silent, for in truth*
>
> *We have no gift to set a statesman right” William Yates*^*10*^

The electrophysiologist, like the poet is a luxury that society may dispense with when times are exceptionally difficult. Our work reduces or resolves troublesome symptoms; in narrow subgroups of the population that we serve, we may increase life expectancy, albeit marginally. In achieving this we expend resources that may be better used elsewhere in difficult times and we bring patients into a healthcare environment that has suddenly become dangerous. We have no expertise that is relevant to the management of an epidemic but we have qualities that might be useful: Being delicate and unnecessary we may serve as an indicator of the health of our surroundings.

The results presented show that in representative centres from several of the worst-affected countries, the arrival of COVID-19 caused a complete cessation of routine electrophysiological intervention. A small number of urgent ablations were performed in patients without infection. Although arrhythmias were encountered in patients hospitalised with COVID-19, these were all either terminal events in the context of cardiomyopathy resulting from the condition or were related to the metabolic and haemodynamic stress arising from infection and mechanical ventilation. No instance was encountered of an arrhythmia arising from COVID-19 requiring urgent ablation.

### COVID-19 and the cardiovascular system

Severe acute respiratory syndrome coronarvirus-2 (SARS-CoV-2), the agent responsible for COVID-19 has an affinity for angiotensin converting enzyme 2 (ACE2) receptors.^11^ This is central to the pathophysiology of the condition, leading to pneumonia and in critical stages, associated fulminant acute respiratory distress syndrome (ARDS) and multi-organ failure.^12^ The primary affected organ is the lung but cardiovascular injury is also common. There is enhanced expression of ACE-2 in those with cardiovascular conditions, possibly accounting for the apparent greater severity of COVID-19 related illness in these patients. In one study, a majority (58%) of those with severe infection had a background of hypetension, and non-specifically 44% had a history of ‘arrhythmia’.^13^

Binding of the virus to ACE2 receptors in the lung and the heart initiates the acute inflammatory response. Pneumonia results, but also evidence of myocarditis; those with a rise in high-sensitivity Troponin I are more likely to require admission to intensive care.^14^ There is a strong association between a history of hypertension and mortality from COVID-19. Systolic blood pressure was significantly higher in those with COVID-19 treated in intensive care units compared to those not requiring this level of care (144 versus 122, P<0.001).^14^ This may relate to up-regulation of ACE2 receptors in those receiving ACE inhibiting drugs.

A history of ischaemic heart disease also predicts mortality in COVID-19. Any acute inflammatory condition can destabilise coronary plaque and triger acute coronary syndromes. A minority of patients with COVID-19 may present acutely with cardiac sounding symptoms.^13^ It is not clear whether the apparent high mortality in patients with ischaemic heart disease represents an effect on ACE2 receptor expression, a vulnerability to ischaemic complications of the systemic illness or a bias against such patients when ventilatiors must be rationed. The pathophysiology of COVID-19 and its routes of systemic transmission remain incompletely understood. Observations had been noted of its neuroinvasive potential via peripheral neuronal trans-synaptic route and viral preponderance in organs with low expression of ACE2.^15^

Brugada syndrome is a concern as the prolonged febrile illness characteristic of COVID-19 has the propensity to trigger arrhythmias. No such case has been reported, but like patients with heart failure and ischaemic heart disease, those with Brugada syndrome should take particular care to isolate themselves from likely sources of infection.

No specific pharmacological therapy exists, but drugs being trialled in treating aspects of COVID-19 include anti-retrovirals and hydrochloroquine. These drugs may cause cardiotoxicitiy and prolong corrected QT intervals. No instance of a life-threatening arrhythmia from this source has been reported.

### Transmission to Healthcare providers

The risk of COVID-19 transmission to healthcare workers is high, with over 8% reported in Italy.^16^ Ophthalmologists have been disproportionately affected, probably because of the close contact with the patients in whom conjunctivitis has been a presenting symptom.^17^ Intensivists are at risk due to their inevitable extensive contact with severely affected patients and due to aerosol generation during intubation. Cardiology, including ablation and trans-esophageal echo also involves aerosol generating procedures (AGP) and close proximity to the patient.

Under normal circumstances, a patient entering the cardiac catheter lab will encounter approximately 10 healthcare professionals before moving to a ward for recovery. The combination of numerous close contacts and fomites creates a risk of transmission. It makes sense to defer procedures whenever possible. Emergency procedures should be undertaken with strict infection control including the use of personal protective equipment (PPE).

Before COVID-19, China was already experienced in infection control protocols due to their experience of Severe Acute Respiratory Syndrome (SARS) in 2002-2003, and a rapid and effective response took place once the problem was recognised. In contrast, without any recent experience to draw on, Italy and the UK had been relatively unprepared for the scale of the events.

Like China, Hong Kong was culturally sensitised by experience of SARS and produced swift and effective intervention to protect the public and healthcare workers, avoiding the spread of infection in healthcare settings. This involved a 3-level hierarchy of control measures: administrative control, environmental control and the use of PPE.^17^ Rigorous screening of the prospective patients took place to avoid bringing the virus into the uninfected healthcare environment.

### EP patient cohort

Many patients scheduled for EP procedures fall into categories at high risk of death if exposed to COVID-19. In the London cohort, the average age is 65, and >60% are male.^18^ Co-existing diabetes, hypertension or heart failure are common. All of these, particularly hypertension are strong risk factors for COVID-19 related mortality. It was therefore obligatory to halt the performance of routine ablation procedures until the risk recedes.

Not all EP patients can wait on indefinitely. Guidelines published by NHS England,^19^ include narrow criteria for ablation in this crisis, recommending that it is limited to cases of rapidly conducted pre-excited atrial fibrillation, heart failure secondary to tachycardia, and ventricular tachycardia that is controllable with medication. All of the ablations conducted in all 3 centres during the period of restricted activity met these criteria (**figure 2, table 2)**.

### Catheter lab protocols

Strict guidelines were put in place in each centre to minimise the risk to patients and staff in cases where ablation had to be performed. Local protocols mirored published consensus documents.^20^ Strict isolation measures were applied before and after any procedure. During the procedure, staff wore PPE.

### EP Industry

The reduction in electrophysiology caseload related to the pandemic will effect the EP industry. Companies which supply equipment have shut offices and distribution networks. Loss of revenue will last for many months. Companies may prosper and may help the fight against the pandemic by switching production to items of relevance to that fight: Ventilators and associated equipment, viral test kits and thermometers.

### Public Health Measures

In China, the official recognition of the seriousness of the outbreak and the take-over by central government of direct responsibility for its handling coincided with the introduction of an integrated set of measures.^7^ Best publicised was the lock-down of society and industry to achieve extreme social distancing. The less-publicised adjunct to the lock-down was the introduction of a system for screening all persons with fever or with a history of contact with known cases, so that they could be isolated in designated facilities. The use of electronic technology and surveillance served as an adjunct to these measures.

The short-term impact of China’s program of public health measures is evident from the progression of the epidemic (**figure 1**). In China, a single decisive act brought all necessary measures into being at a time when deaths were in low double figures. In Italy and in the UK as across Europe, distancing measures were introduced piecemeal and later in the course of events, and the infrastructure for the identification of infected individuals and the tracing, testing and isolation of contacts has not yet been developed.

The experience from Wenzhou shows that with energetic public health intervention, the epidemic may be suppressed, and normal life may resume with precautions. In the longer term, a vaccine may permit us to dispense with these precautions. The less stringent interventions chosen by the UK and other North European countries risk allowing a lingering epidemic that drags on for so long that by the time a vaccine comes, production and supply chains will have fallen into habitual disuse and operators will have lost familiarity with their craft. Many patients will also have died by then.

## CONCLUSION

In the Spring of 2020, as in the Summer of 1914, it feels as though “the lamps are going out all over Europe”, but there is reason for hope. Vigorous public health measures have suppressed the epidemic in parts of Asia, permitting a resumption of some normal routine activity with stringent precautions.

## Data Availability

Data in the manuscript is fully available from all of the co-authors.

## REFERENCES

1. Lai CC, Shih TP, Ko WC, Tang HJ, Hsueh PR. Severe acute respiratory syndrome coronavirus 2 (SARS-CoV-2) and coronarvirus disease-2019 (COVID-19): The epidemic and the challenges. Int J Antimicrob Agents. 2020. 55(3):105924. doi: 10.1016/j.ijantimicag.2020.105924

2. Wang C, Horby PW, Hayden FG, Gao GF. A novel coronavirus outbreak of global health concern. Lancet 2020. 395 (10223); 470–473. https://doi.org/10.1016/s0140-6736(20)30185-9.

3. Chen C, Zhou Y, Wang DW. Sars-cOv-2: a potential novel etiology of fulminant myocarditis. Herz. 2020. https://doi.org/10.1007/s00059-020-04909-z.

4. Hu H, Ma F, Wei X, Fang Y. Coronarvirus Fulminant Myocarditis Saved With Glucocorticoid and Human Immunoglobulin. Eur Heart J. 2020. Ehaa190, htpps://doi.org/10.1093/eurheartj/ehaa190.

5. Driggin E, Madhavan MV, Bikdeli B, Chuich T, Laracy J, Bondi-Zoccai G et al. Cardiovascular Considerations for Patients, Health Care Workers, and Health Systems During the Coronarvirus Disease 2019 (COVID-19) Pandemic. J Am Coll Cardiol. 2020. S0735-1097(20)34637-4. doi: 10.1016/j.jacc.2020.03.031.

6. Clinical findings in a group of patients infected with the 2019 novel coronarvirus (SARS-CoV-2) outside of Wuhan, China: a retrospective case series. BMJ 2020; 368:M6060. https://doi.org/10.1136/bmj.m606.

7. Gong F, Xiong Y, Ziao J, Lin L, Liu X, Wang D et al. China’s local governments are combating COVID-19 with unprecedented responses-from a Wenzhou governance perspective. Front Med 2020. doi: 10.1007/s11684-020-0755-z.

8. Spinelli A, Pellino G. COVID-19 pandemic: perspectives on an unfolding crisis. BJS 2020. https://doi.org/10.1002/bjs.11627

9. World Health Organisation (WHO) coronarvirus disease (COVID-19) outbreak https://experience.arcgis.com/experience/685dace521648f8a5beeeee1b9125cd

10. Yates WB (1916). “A reason for keeping silent.” In: Edith Wharton, The Book of the Homeless. 1916. Macmillan.

11. Lu R, Zhao X, Li J, Niu P, Yang B, Wu H et al. Genomic characterisation and epidemiology of 2019 novel coronavirus: implications for virus orgins and receptor binding. Lancet 2020; 395(10224):565–574.

12. Chen N, Zhou M, Dong X, Qu J, Gong F, Han Y et al. Epidemiological and clinical characteristics of 99 cases of 2019 novel coronarvirus pneumonia in Wuhan, China: a descriptive study. Lancet. 2020. 395(10223): 507–513. doi: 10.1016/s0140-6736(20)30211-7.

13. Zheng Y, Ma Y, Zhang J, Xiang X. COVID-19 and the cardiovascular system. Nat Rev Cardiol. 2020. https://doi.org/10.1038/s41569-020-0360-5.

14. Huang C, Wang Y, Li X, Ren L, Zhao J, Hu Y et al. Clinical features of patients infected with 2019 novel coronavirus in Wuhan, China. Lancet 2020. doi: https://doi.org/10.1016/S0140-6736(20)30183-5

15. Li Y, Bai W, Hashikawa T. The neuroinvasive potential of SARS-CoV2 may play a role in respiratory failure of COVID-19 patients. J Med Virol 2020; 1–4. doi: 10.1002/jmv.25728.

16. https://www.msf.org/covid-19-urgent-help-needed-across-european-borders-rotect-medical-staff (Date of access: 24.03.2020).

17. Lai T, Tang E, Chau S, Fung K, Li K. Stepping up infection control measures in ophthalmology during the novel coronavirus outbreak: an experience from Hong Kong. Graefe’s Archive for Clinical and Experimental Ophthalmology. https://doi.org/10.1007/s00417-020-04641-8.

18. Leung LW, Bajpai A, Zuberi Z, Li A, Norman M, Kaba R et al. Improving Esophageal Protection during AF ablation: the IMPACT study. Preprint. DOI: 10.1101/2020.01.30.20019158.

19. NHS England Publication; 001559 version 1. Clinical guide for the management of cardiology patients during the coronavirus pandemic. 2020; 1–5. specialty-guide-cardiolgy-coronavirus-v1-20-march.pdf.

20. Welt F, Shah P, Aronow H, Bortnick A, Henry T, Sherwood M et al. Catheterization Laboratory Considerations During the Coronavirus (COVID-19) Pandemic: From ACC’s Interventional Council and SCAI. J Am Coll Cardiol. 2020. Doi 10.1016/j.jacc.2020.03.021.

21. Viscount Grey of Fallodon: Twenty-Five Years 1892–1916. Frederick A. Stokes,New York, 1925.

